# Modelling the impact of population mobility, post-infection immunity and vaccination on SARS-CoV-2 transmission in the Dominican Republic

**DOI:** 10.1101/2023.10.05.23296586

**Authors:** Emilie Finch, Eric J Nilles, Cecilia Then Paulino, Ronald Skewes-Ramm, Colleen Lau, Rachel Lowe, Adam J Kucharski

## Abstract

COVID-19 epidemic dynamics are driven by a complex interplay of factors including population behaviour, government interventions, new variants, vaccination campaigns and immunity from prior infections. We aimed to quantify the epidemic drivers of SARS-CoV-2 dynamics in the Dominican Republic, an upper-middle income country of 10.8 million people, and assess the impact of the vaccination campaign implemented in February 2021 in saving lives and averting hospitalisations.

We used an age-structured, multi-variant transmission dynamic model to characterise epidemic drivers in the Dominican Republic and explore counterfactual scenarios around vaccination coverage and population mobility. We fit the model to reported deaths, hospital bed occupancy, ICU bed occupancy and seroprevalence data until December 2021 and simulated epidemic trajectories under different counterfactual vaccination scenarios.

We estimate that vaccination averted 5040 hospital admissions (95% CrI: 4750 - 5350), 1500 ICU admissions (95% CrI: 1420 - 1590) and 544 deaths (95% CrI: 488 - 606) in the first 6 months of the campaign. We also found that early vaccination with Sinovac-CoronaVac was preferable to delayed vaccination using a product with higher efficacy. We investigated the trade-off between changes in vaccination coverage and population mobility to understand how much relaxation of social distancing measures vaccination was able to ‘buy’ in the later stages of a pandemic. We found that if no vaccination had occurred, an additional decrease of 10-20% in population mobility would have been required to maintain the same death and hospitalisation outcomes. We found SARS-CoV-2 transmission dynamics in the Dominican Republic were driven by substantial accumulation of immunity during the first two years of the pandemic but that, despite this, vaccination was essential in enabling a return to pre-pandemic mobility levels without incurring considerable additional morbidity and mortality.

## Introduction

During 2020-22, many countries experienced a significant burden of COVID-19 and imposed non-pharmaceutical interventions (NPIs) aiming to control SARS-CoV-2 transmission. Despite this, countries experienced markedly different epidemic dynamics, mediated by a complex interplay of factors including population behaviour, government interventions, the introduction of new variants, the roll-out of vaccination campaigns and prior levels of transmission. Serological surveys have proven crucial to understand global and national landscapes of population immunity, as well as indicating the extent of prior exposure to SARS-CoV-2 (1). Some countries pursued an elimination strategy throughout the pre-Omicron era, with stringent public health interventions resulting in low levels of seroprevalence towards the end of 2021. For example, in Hong Kong, serosurveillance studies found <1% of sera tested was positive for anti-N SARS-CoV-2 IgG prior to March 2021 (2). Other countries, such as the UK and many EU countries, saw epidemic waves linked to the strengthening and relaxing of public health interventions, alongside the emergence of more transmissible variants. In England, approximately 20% of the population were estimated to have been infected with SARS-CoV-2 by July 2021 (3). In contrast, SARS-CoV-2 transmission was largely unmitigated in some settings such as in Manaus, Brazil, where 76% of the population are thought to have been infected by October 2020 (4).

Latin America and the Caribbean was a global hotspot for SARS-CoV-2 transmission during 2020-21, prior to the emergence of the Omicron variant, which caused large epidemics globally (5,6). The Dominican Republic is an upper-middle income country with a population of 10.8 million, which shares the island of Hispañola in the Caribbean with Haiti. They reported their first case of COVID-19 on the 1st of March 2020, which was followed by the imposition of strict public health measures. These began to be relaxed in July 2021 and were mostly lifted with the reopening of schools and relaxation of curfew measures in October 2021. In the first two years of the pandemic, the Dominican Republic experienced four waves of transmission: the first peaked in August 2020, with cases increasing again in November 2020 before a second peak in January 2021. A third wave of transmission took place over the summer of 2021, following the introduction of more transmissible variants, including Mu, with cases rising sharply to peak in July 2021. Following the introduction of the Delta variant, a fourth wave took place in October and November 2021. A nationally representative serological survey involving 6683 individuals from 3832 households took place between June and September 2021 (7). Results from the serological survey estimated that 76.6% (95% CI 70.1 – 82.5) of the population had been previously infected by the study midpoint. This vastly exceeded earlier estimates constructed from coarse reported data for the Dominican Republic, and the wider region of Latin America and the Caribbean (8).

The government of the Dominican Republic launched a national COVID-19 vaccination programme on 16th February 2021 initially focusing on health-care professionals and then following a three-phase age-based approach. Booster vaccination for highly vulnerable individuals began in July 2021, with the Dominican Republic being the first country in the Americas to approve vaccination with a third dose.

Approximately 90% of vaccine doses administered were Sinovac-CoronaVac (an inactivated viral vaccine), with Oxford/AstraZeneca vaccine (ChAdOx1-S, an adenovirus vector vaccine) and Pfizer/BioNTech (BNT162b2, mRNA vaccine) also administered (7). By the current study’s endpoint (15th December 2021), 62% of the population had received at least one dose of a COVID-19 vaccine and 50% had received two doses.

Mathematical models have been used throughout the pandemic to provide decision-support to policy makers through estimation of key epidemiological parameters, forecasts of future incidence, projections of epidemic trajectories under different scenarios, and quantification of the impact of non-pharmaceutical interventions. However, despite regular and in-depth modelling decision-support for high-income countries, there has been a lack of equivalent modelling analysis to understand transmission and control in low-and middle-income countries (9–15). To address this gap, we used an age-structured transmission dynamic model to quantify the drivers of epidemic dynamics in the Dominican Republic during the first two years of the pandemic, and to assess the impact of the vaccination campaign on COVID-19 hospitalisations and deaths.

## Methods

### Data

For this analysis we incorporated multiple data streams. Aggregated daily reported deaths were collected from the Dominican Republic’s COVID-19 Dashboard. Aggregated daily hospital and ICU bed occupancy were scraped from daily COVID-19 bulletins published by the Ministerio de Salud Pública y Asistencia Social, available from 19th September 2021 onward (17). We also used serological data from a nationally representative SARS-CoV-2 seroprevalence survey undertaken between June -October 2021. Further details of survey methodology and findings are available elsewhere (7).

We used data on the daily number of second vaccine doses distributed in the population (18). To obtain estimates of age-stratified vaccination rates, we assumed that daily doses were evenly distributed between eligible age groups as per the government’s vaccination program (Supplementary Table S.9). If an eligible age group became fully vaccinated during the vaccination allocation, remaining vaccine doses were distributed between the remaining eligible age groups or, if all were fully vaccinated, between adults > 20, mimicking the vaccination of younger health care workers or those with chronic health conditions. Estimated vaccination coverage by age group over time is shown in Figure 2.

**Figure 1:**
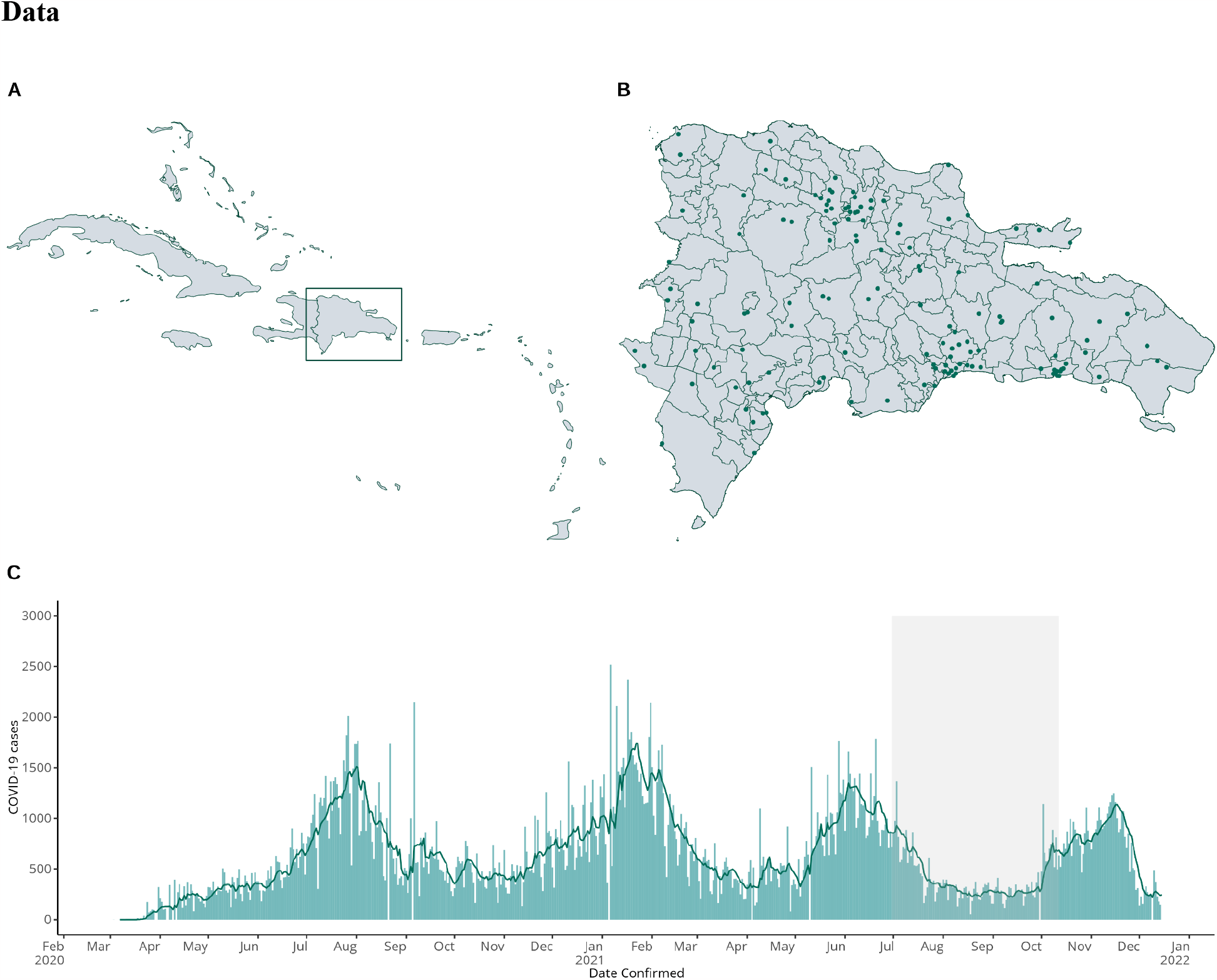
Map of the study setting and time-series of COVID-19 cases. Figure showing a map of the Caribbean with the location of the Dominican Republic shown in a box (A), a map of the Dominican Republic showing clusters sampled in the 2021 serosurvey (B), and daily COVID-19 cases in the Dominican Republic (bars) and the 7-day moving average (line) from March 2020 – January 2022 (C). The shaded grey area indicates the timing of the serological survey.

**Figure 2:**
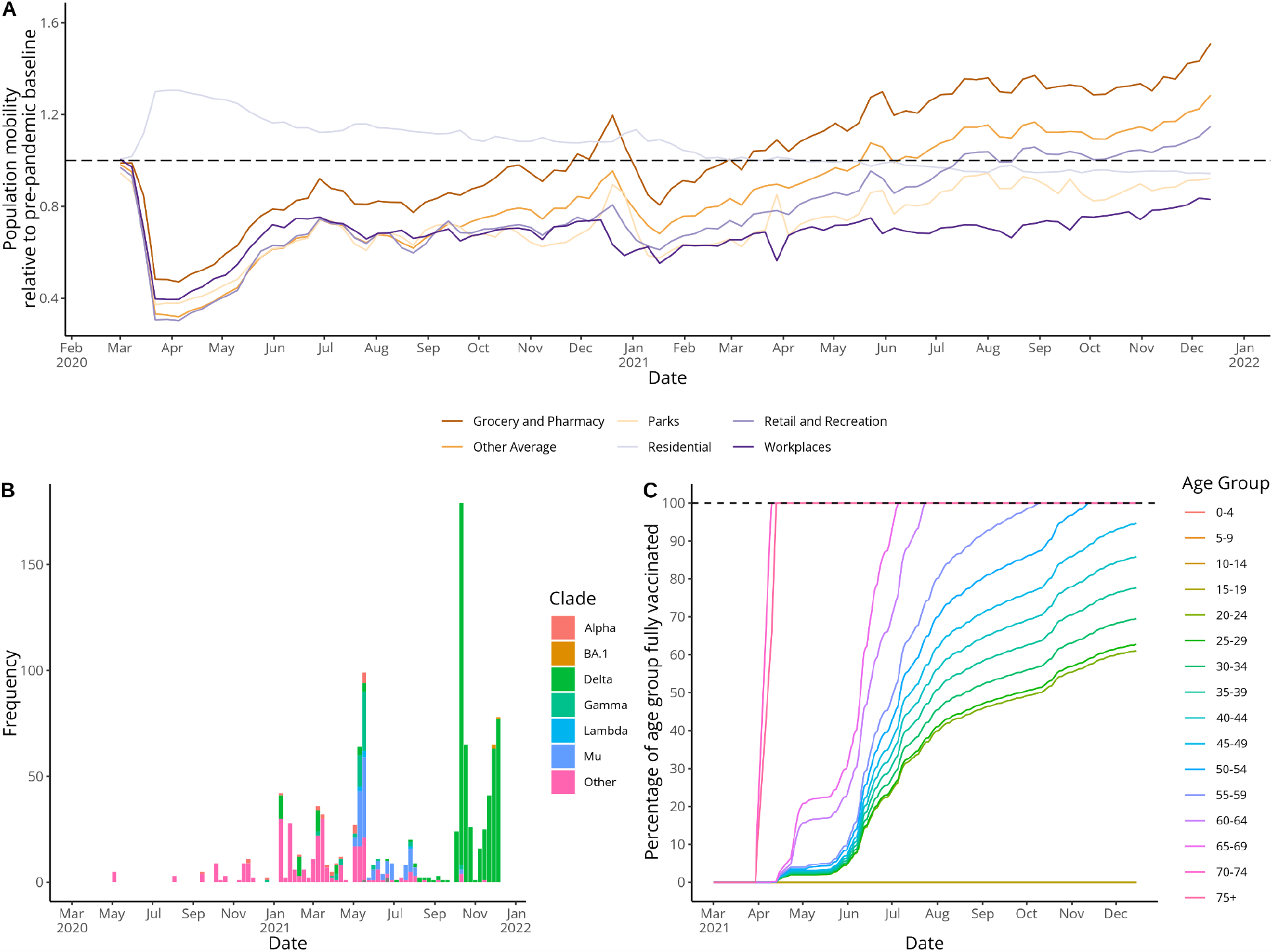
Google mobility data, SARS-CoV-2 sequence data, and estimated vaccination coverage by age. Panel A shows Google mobility data showing the proportional change in population mobility in different locations relative to a pre-pandemic baseline, panel B shows the frequency of SARS-CoV-2 sequences from the Dominican Republic on GISAID by variant and panel C shows estimated vaccination coverage by age assuming vaccine doses were evenly distributed between eligible age groups.

### Transmission dynamic model

We used covidM, an age-stratified, deterministic, compartmental model originally developed to model the effects of NPIs on SARS-CoV-2 transmission in the UK, and described fully elsewhere (19,24,25). In brief, covidM is structured into 5-year age groups, with individuals moving from a susceptible state (S) to an exposed state (E) and then either to a pre-clinical and clinical infected state (I_p_ followed by I_c_) or a sub-clinical infected state (I_s_) and finally to a recovered state (R). The model explicitly considers two variants of SARS-CoV-2; wild-type and B.1.617.2 (Delta), while the introduction of other variants in the Dominican Republic in the intervening period (which include Mu, Gamma and Alpha) is captured through a gradual increase in transmissibility in the first half of 2021, following a logistic function. We fit the model to daily reported deaths, daily hospital and ICU bed occupancy and a cross-sectional seroprevalence estimate from June -October 2021. We fit the model using data until 15th December 2021, when cases began to increase due to the Omicron variant, and only simulate epidemic trajectories until this point.

Hospitalisation, ICU admission and death are modelled as observation processes according to age-specific infection-severe ratios, infection-critical ratios and infection-fatality ratios based on estimates from the literature. These are adjusted on the log odds scale by several fitted parameters and delays from infection to hospitalisation, ICU admission and death are estimated during the model fitting process. We assume that the observed number of deaths, hospital bed occupancy and ICU bed occupancy are distributed according to a negative binomial distribution, with the overdispersion parameter estimated during the model fitting process. We used a skew-normal likelihood for seroprevalence with the same mean and 95% confidence interval as reported for the data evaluated for the period of the serosurvey.

We consider a central waning assumption corresponding to 15% loss of post-infection protection after 1 year, and conduct sensitivity analysis around different waning assumptions (Supplementary Tables S.6 and S.7). Full details on model equations, fixed and fitted model parameters can be found in Supplementary Tables S1-3.

### Vaccination parameters

We also incorporated information on SARS-CoV-2 vaccination in the Dominican Republic, using data collated by Our World in Data (10). Fully vaccinated individuals moved to a vaccinated model compartment (V) from which, subject to vaccine waning parameters, they can move to the exposed state (E) or directly to a sub-clinical infection (I_s_). We assume vaccinated individuals have a lower probability of clinical or sub-clinical infection but that, once infected, they have the same infectiousness as non-vaccinated individuals. We model vaccine efficacy against infection (*vei*), which determines the probability that a vaccinated individual enters the Exposed compartment, and vaccine efficacy against disease given infection (*ved*|*i*), which determines the probability that a vaccinated individual develops sub-clinical or asymptomatic disease directly (see Figure 3).

**Figure 3:**
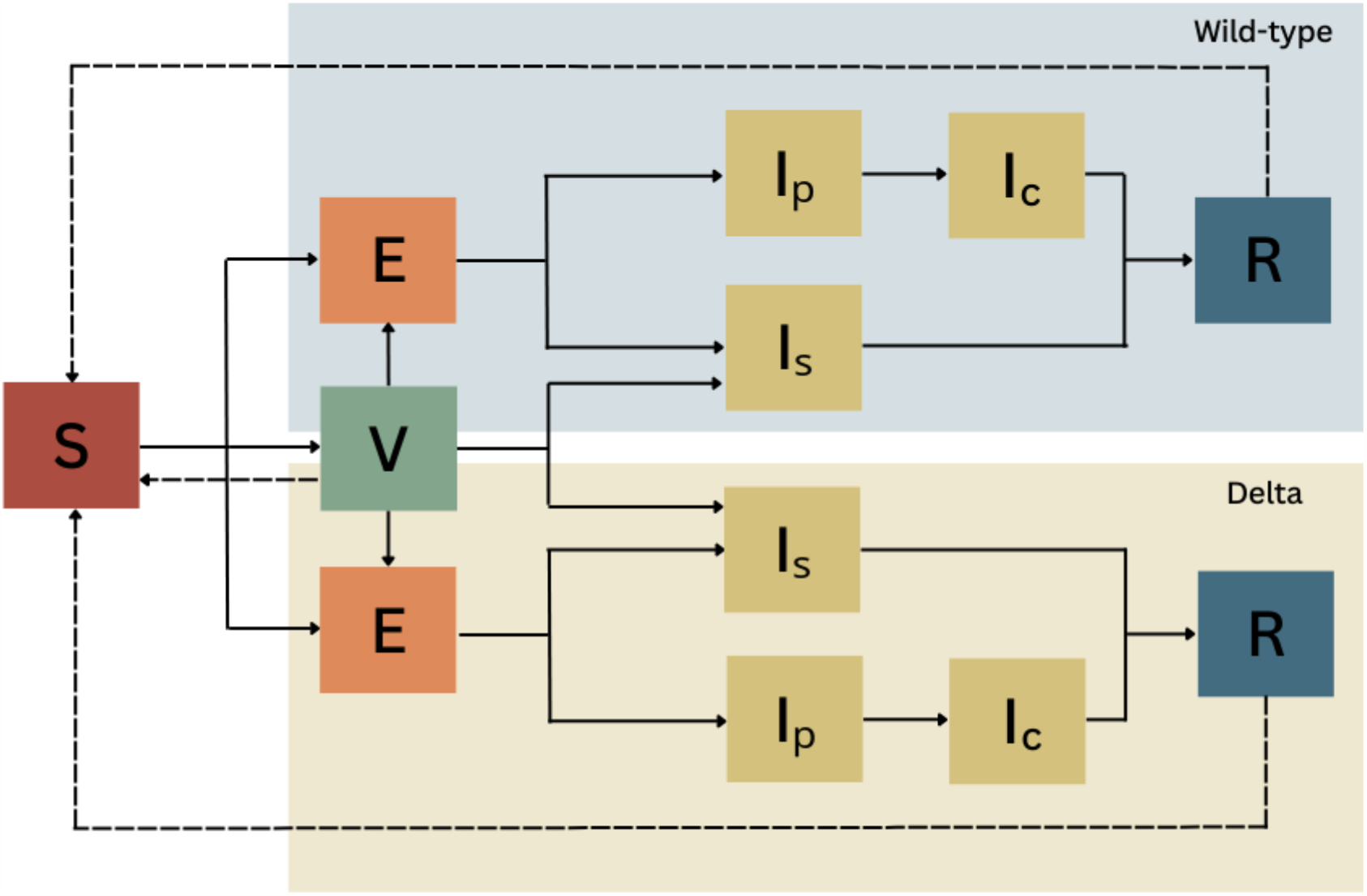
Model schematic showing a two-variant SEIR model structure with a vaccination compartment (V).

As 90% of the primary course of vaccinations given in the Dominican Republic were CoronaVac (Sinovac COVID-19 vaccine) we used vaccination efficacy parameters based on the literature available for this vaccine product. We model differing vaccine efficacy by strain but assume the same vaccine efficacy across age groups. Note that we do not directly model vaccine efficacy against hospitalisation or death.

### Mobility

To estimate changes in population behaviour during the pandemic we used population mobility captured by Google’s COVID-19 Community Mobility Reports as a proxy. We mapped changes in mobility to changes in contact rates using the relationship between mobility and contact survey data found in the UK (19,41). The representativeness of Google mobility data is dependent on the proportion of the population with smartphones using Google products. As this differs substantially between the UK and the Dominican Republic, we infer a weighting between UK-adjusted contact rates and pre-pandemic baseline contact rates in the Dominican Republic, fitting a separate weighting parameter for each year of the simulation period (35). We found the UK-adjusted value was given more weight in the first year of the pandemic than the second, suggesting that the relationship between measured population mobility and contact rates changed during the pandemic. We considered four categories of contacts: home, work, school and other. School contact rates were set to zero during school closures and school holiday periods.

### Model fitting

We performed Bayesian inference using Markov chain Monte Carlo to estimate model parameters. We used the Differential Evolution Markov Chain Monte Carlo (DE-MCMC) algorithm which combines a genetic algorithm (Differential Evolution) with MCMC (26). Here, multiple Markov chains are run in parallel and learn from one another to determine the scale and orientation of the proposal distribution, which allows for more efficient exploration of a complex parameter space than traditional Metropolis-Hastings algorithms, particularly when considering correlated parameters. Model convergence was assessed using trace plots of MCMC chains and the Rhat statistic (27).

### Counterfactual analysis

We conducted counterfactual scenario analysis using the fitted model to simulate epidemic trajectories under different scenarios. We considered five key scenarios with changes applied from the beginning of the vaccination campaign (15th February 2021) until the end of the analysis period.

For Scenarios 3 and 5 a vaccination programme using a Pfizer/BioNTech or Oxford/AstraZeneca efficacy profile with a delay of two months is considered. Here vaccination would begin 15th April 2021, aligning with initial deliveries of Oxford/AstraZeneca vaccine doses through COVAX (42).

For each scenario we ran 500 simulations, drawing parameters from the posterior distribution of each fitted parameter, and calculated the difference in hospital admissions, ICU admissions and deaths from the original model fit to estimate the impact of the scenario considered.

We also conducted a counterfactual analysis to examine the trade-off between differing levels of vaccination coverage and changes in population mobility on deaths, hospital admissions and ICU admissions during the time period between 16th February -16th August 2021 . We considered 11 vaccination coverage scenarios with coverage by 16th August 2021 ranging from 0 to 100% in 10% increments, and 9 population mobility scenarios changing ‘Work’ and ‘Other’ mobility by an extra -40% to +40% compared with the Google mobility data during the simulation period. Vaccination allocation for each scenario was performed in a similar way as described above, where daily vaccinations are multiplied by a factor equal to 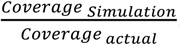. Vaccination doses are then distributed between eligible age groups and (once these are fully vaccinated) between age groups > 20 and then finally between age groups < 20. We then used the fitted model to simulate epidemic trajectories for 99 scenarios, considering all combinations of vaccination coverage and population mobility change, and estimated the impact of the scenario considered as above.

## Results

### COVID-19 transmission dynamics between 2020-2022

Our modelling analysis suggests that, after an initial decline in transmission following a sharp reduction in social interactions, COVID-19 dynamics were driven by substantial accumulation of immunity throughout 2020-2022 (Figure 4), as well as the spread of novel variants such as Mu in mid-2021 and Delta in late 2021. By jointly fitting to reported deaths, hospital bed occupancy, ICU bed occupancy and seroprevalence data, the model reproduced the overall observed epidemic dynamics. Estimated deaths did not track closely with observed deaths during the first wave from March -September 2020, and deaths and hospitalisations were slightly underestimated during the second wave in January 2021.

**Figure 4:**
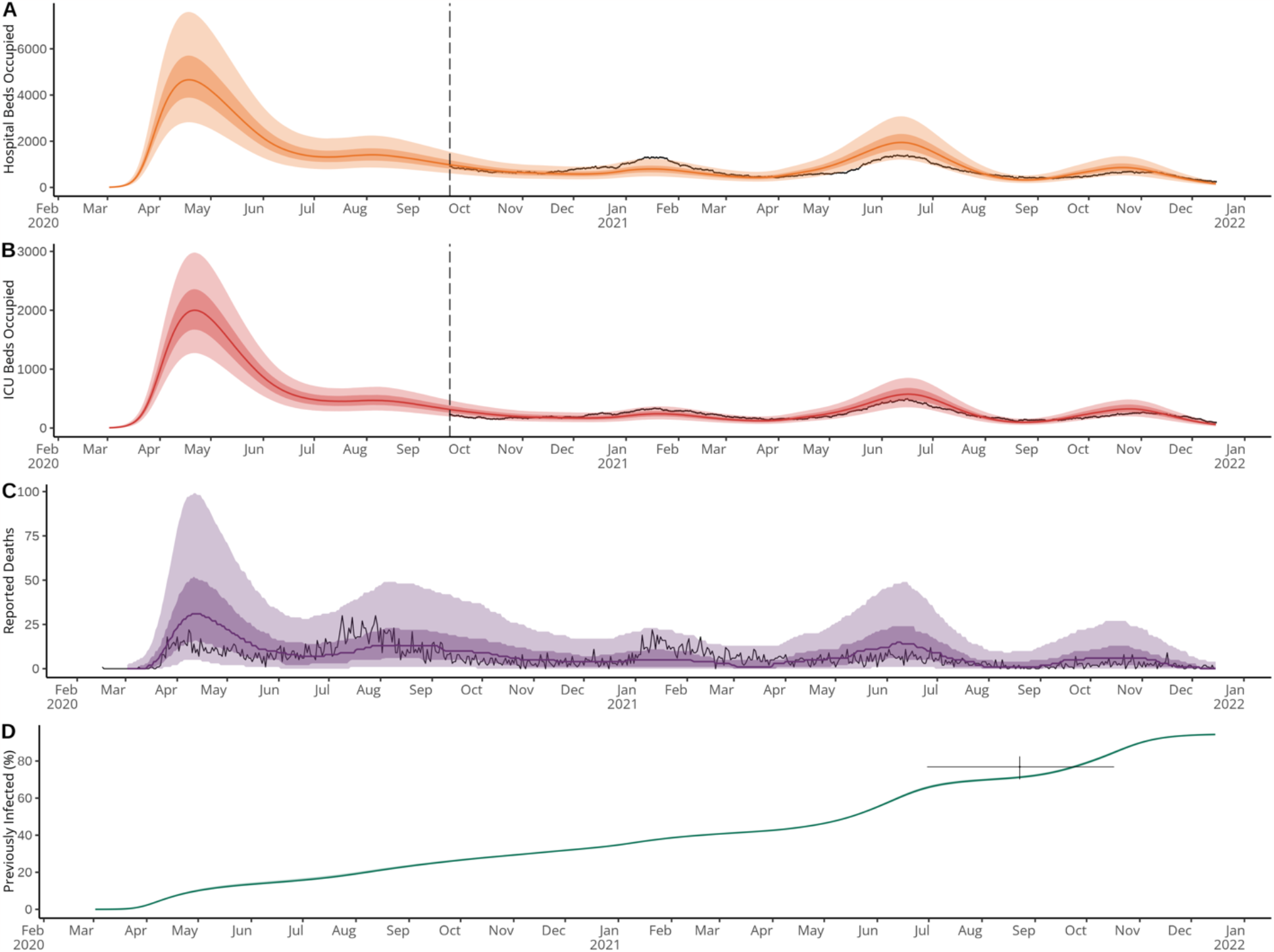
Comparison of model fit to observed data in the Dominican Republic from February 2020 to December 2021. Panels show comparison between modelled and observed hospital bed occupancy (A), ICU bed occupancy (B), reported deaths (C) and proportion of the population previously infected (D). Black lines show observed data, with horizontal dashed lines in panels (A) and (B) indicating the point at which hospitalisation data became available. For panel (B) the black cross shows the duration of the serosurvey (horizontal line) and the 95% confidence interval around the central estimate (vertical line). Modelled hospital bed occupancy, ICU bed occupancy, deaths, and proportion previously infected, are shown in orange, red, purple and green respectively, with associated 50% and 95% credible intervals in surrounding ribbons. Note that uncertainty in the observation process is included in modelled outputs for surveillance data streams (A, B and C) but not for the proportion previously infected (D).

Reconstructing the underlying epidemic dynamics, we found that changes in the effective reproduction number, R_t_, reflected changes in contact rates derived from Google mobility data during 2020, but were less strongly associated with contact rates during 2021 (Figure 5). We estimated that 33.3% (95% CrI: 33.3-33.4) of the population had been infected by the end of 2020 (Figure 5), ranging from around 45% in those aged 20-39 to around 20% in those aged under 19 and above 70 (Figure S.3). This accumulation of population immunity contributed to a decline in transmission, with R_t_ remaining around 1 despite gradual increases in contact rates from May 2020. By the end of 2021, we estimated that 83.0% (95% CrI: 82.7 - 83.2%) of the population had been infected, ranging from above 90% in those aged 20-39 to around 55% in those over 70 (Figure S.3). Again, high levels of post-infection immunity resulted in reduced levels of transmission despite contact rates approaching pre-pandemic baseline levels, except during the emergence of more transmissible variants in May and September 2021.

**Figure 5:**
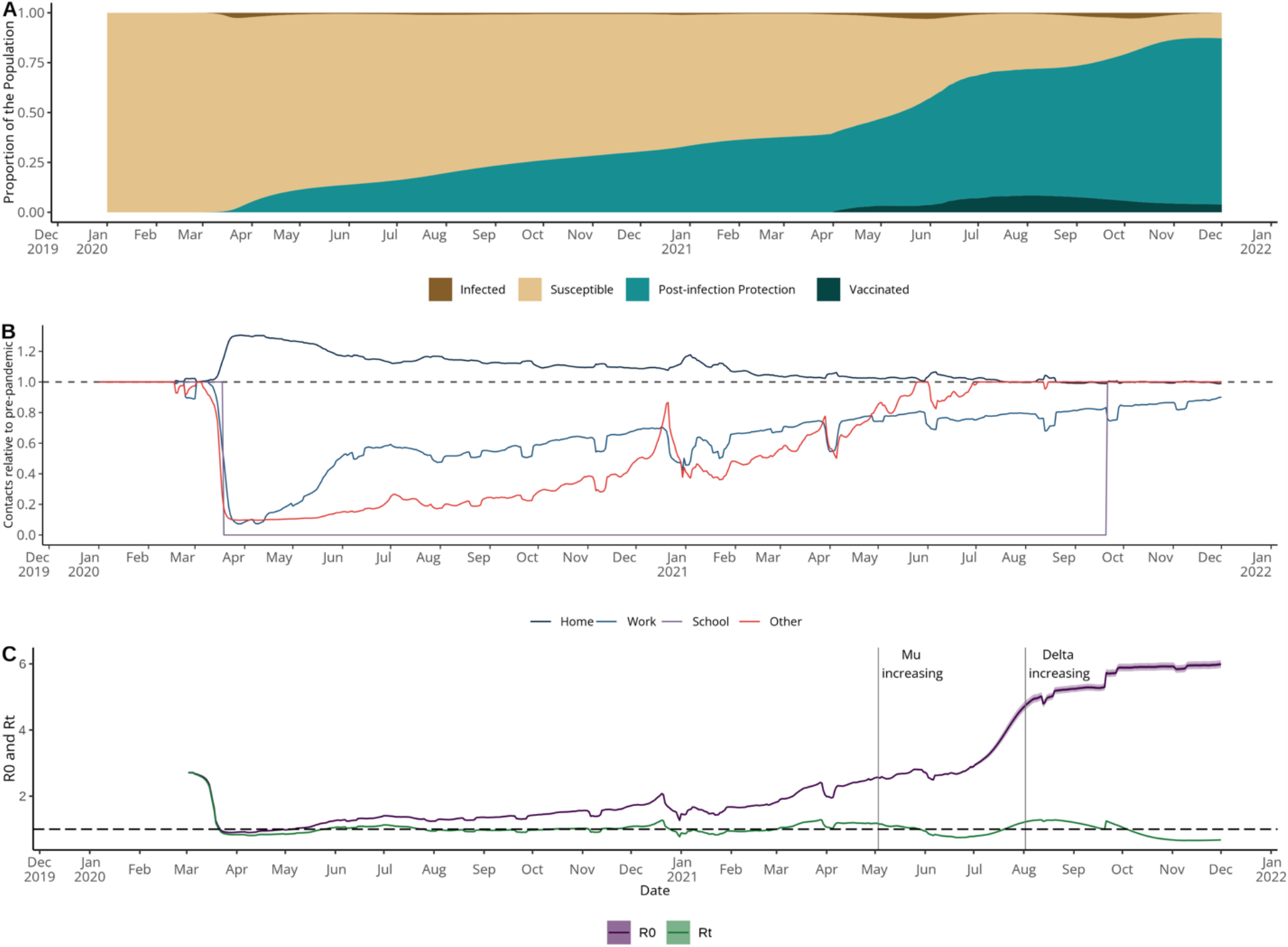
Immune status, contact rates and reproduction number estimates from January 2020 to December 2021. Panel A shows the modelled distribution of immune states in the Dominican Republic over time showing the proportion of the population that are: currently infected (brown), susceptible (beige), protected post-infection (blue) and protected post-vaccination (dark blue). Note that the vaccinated area (dark blue) does not include individuals that were vaccinated post-infection and so does not correspond with observed vaccination coverage. Panel B shows inferred contact rates in home, work, school and other settings relative to a pre-pandemic baseline. Panel C shows estimated R_0_ and R_t_.The vertical lines on Panel C show the time at which Mu sequences start to increase, according to GISAID, the global data science initiative (16).

### Impact of vaccination campaign

To estimate the impact of vaccination, we used our calibrated model to simulate counterfactual epidemic trajectories under different scenarios. First, we considered a ‘no vaccination scenario’, estimating deaths, hospital and ICU admissions in the absence of any vaccination during 2021 (Figure 6). By comparing these counterfactual outcomes to the original model estimates we were able to estimate the burden averted by the vaccination campaign in the 6 and 10 months following its launch in February 2021 (Table 2). It should be noted that this analysis assumes all other factors remain constant and, in particular, we assume the same changes in contact rates and variant transmissibility as in the original model fitting displayed in Figure 4. This is a simplifying assumption, as in reality it is possible countries would respond to rising COVID-19 cases with impositions of further restrictions or would see accompanying changes in population behaviour.

**Table 1:**
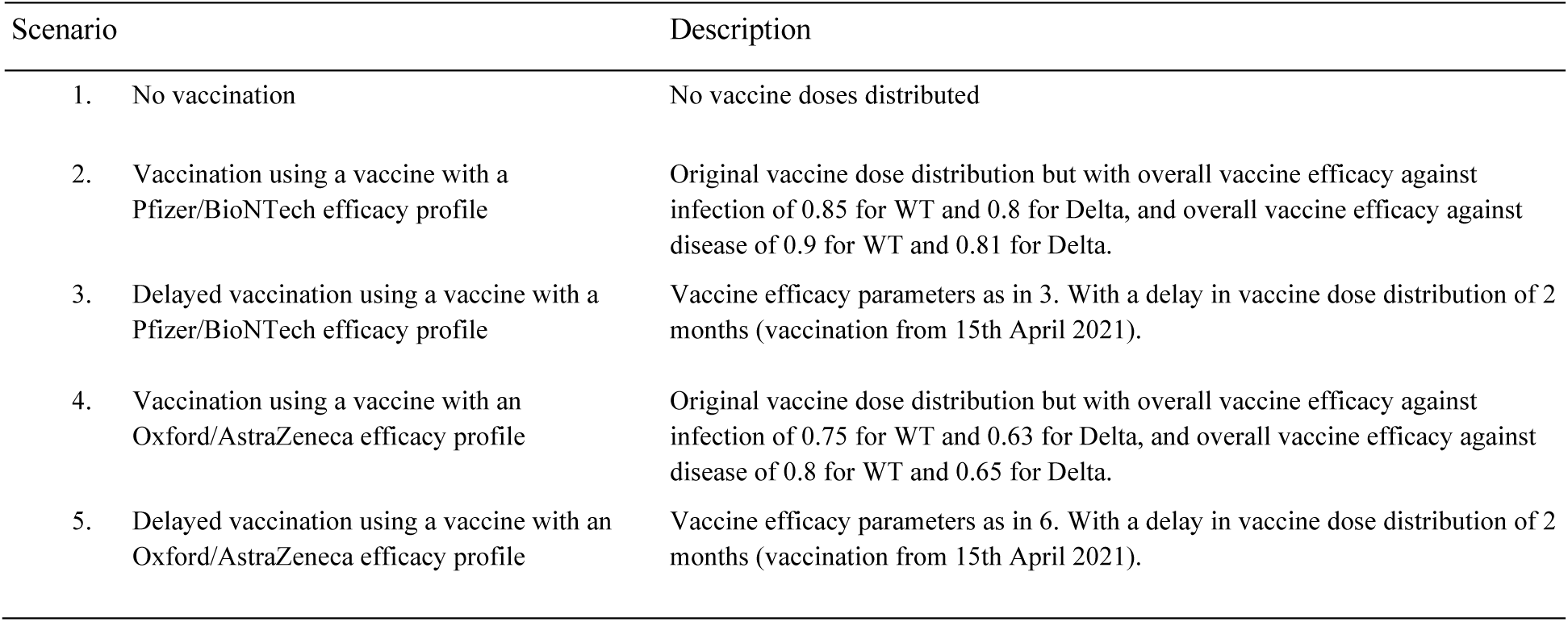
Counterfactual scenarios.

| Scenario | Description |
| --- | --- |
| 1. No vaccination | No vaccine doses distributed |
| 2. Vaccination using a vaccine with a Pfizer/BioNTech efficacy profile | Original vaccine dose distribution but with overall vaccine efficacy against infection of 0.85 for WT and 0.8 for Delta, and overall vaccine efficacy against disease of 0.9 for WT and 0.81 for Delta. |
| 3. Delayed vaccination using a vaccine with a Pfizer/BioNTech efficacy profile | Vaccine efficacy parameters as in 3. With a delay in vaccine dose distribution of 2 months (vaccination from 15th April 2021). |
| 4. Vaccination using a vaccine with an Oxford/AstraZeneca efficacy profile | Original vaccine dose distribution but with overall vaccine efficacy against infection of 0.75 for WT and 0.63 for Delta, and overall vaccine efficacy against disease of 0.8 for WT and 0.65 for Delta. |
| 5. Delayed vaccination using a vaccine with an Oxford/AstraZeneca efficacy profile | Vaccine efficacy parameters as in 6. With a delay in vaccine dose distribution of 2 months (vaccination from 15th April 2021). |

**Table 2:** Estimated total hospital admissions, ICU admissions and deaths under different counterfactual vaccination scenarios. Median values and 95% CrIs are shown from 500 simulations. Estimates are split into the cumulative burden estimated during the 6 months following the beginning of the vaccination campaign (until 16th August 2021, aligning with the Mu wave) and the 10 months following the beginning of the vaccination campaign (until 15th December 2021, aligning with the Mu and Delta waves).

| Scenario | Additional hospital admissions in next 6 months | Additional hospital admissions in next 10 months | Additional ICU admissions in next 6 months | Additional ICU admissions in next 10 months | Additional deaths in next 6 months | Additional deaths in next 10 months |
| --- | --- | --- | --- | --- | --- | --- |
| No vaccination | 5040 (4750 – 5350) | 5150 (4770 – 5540) | 1500 (1420 – 1590) | 1530 (1420 – 1640) | 544 (488 – 606) | 554 (487 – 627) |
| Pfizer efficacy or equivalent | -1830 (-1940 – -1730) | -3410 (-3790 – -3070) | -542 (-573 – -512) | -1020 (-1120 – -921) | -193 (-215 – -174) | -340 (-414 – -273) |
| Pfizer efficacy or equivalent and delay | 2980 (2760 – 3210) | 124 (-364 – 596) | 811 (753 – 870) | -65 (-188 – 60) | 283 (236 – 332) | 17 (-89 – 114) |
| AZ efficacy or equivalent | -1040 (-1100 – -981) | -1920 (-2150 – -1700) | -307 (-325 – -290) | -577 (-642 – -515) | -109 (-122 – -98) | -193 (-240 – -150) |
| AZ efficacy or equivalent and delay | 3220 (3000 – 3450) | 1090 (654 – 1520) | 892 (834 – 952) | 234 (124 – 348) | 313 (267 – 361) | 113 (20 – 200) |

**Figure 6:**
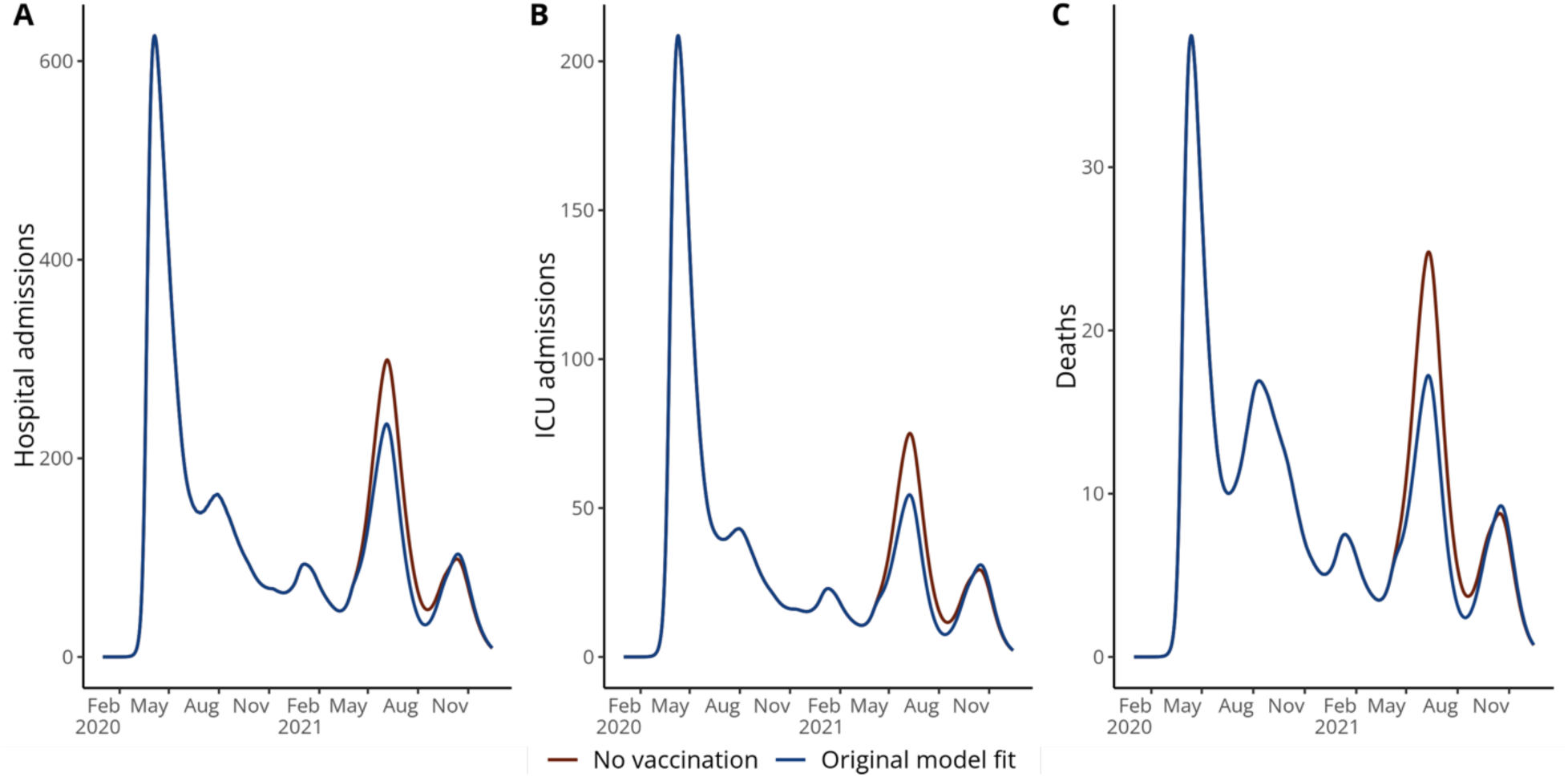
Impact of vaccination campaign. Figure showing modelled deaths (A), hospital admissions (B) and ICU admissions (C) from the original model fit (blue) and from a ‘no vaccination’ counterfactual (red). Lines show the median value from 500 simulations. To facilitate comparison between scenarios, modelled deaths do not include uncertainty generated through the observation process and are therefore higher than those shown in the model fit in Figure 4.

We estimated that the vaccination campaign averted 5040 hospital admissions (95% CrI: 4750 - 5350), 1500 ICU admissions (95% CrI: 1420 - 1590) and 544 deaths (95% CrI: 488 - 606) in the 6 months following its launch. This is equivalent to averting 20.3% (95% CrI: 15.9 - 24.3) of hospital admissions, 24.9% (95% CrI: 20.8 - 28.7) of ICU admissions and 27.2% (95% CrI: 19.5 - 34.1) of reported deaths considering the median values expected under a ‘no vaccination’ scenario. Notably, under a ‘no vaccination scenario’, we estimated ICU capacity would have been exceeded and hospital bed capacity almost reached, given population behaviour and variant introductions observed in 2021 (17).

Given the challenges and inequities of vaccine availability in real-time, with some products available at scale before others, we evaluated the impact of using alternative vaccine products with or without a delay in the vaccination programme on deaths, hospital and ICU admissions.

We estimated that while vaccination with a more efficacious product would have reduced hospitalisations, ICU admissions and deaths, delaying the vaccination campaign to vaccinate with a more efficacious product would have resulted in a higher overall burden in subsequent waves in 2021 (Figure 7). This is due both to the speed of vaccination rollout, with 50% of the population receiving a two-dose primary series by the end of 2021, as well as the introduction of variants of concern or interest (particularly the Mu variant) in the summer of 2021 (18).

**Figure 7:**
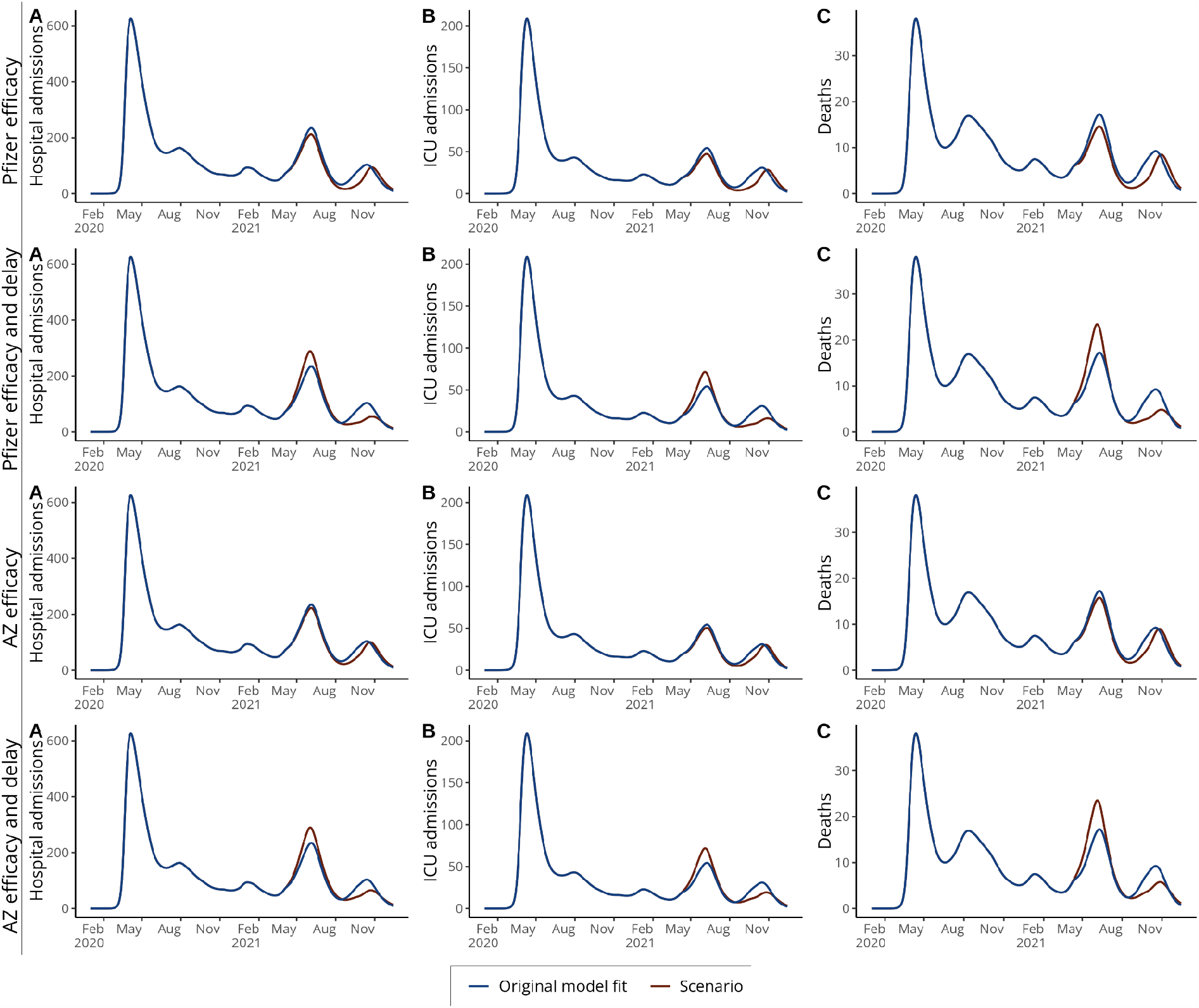
Impact of vaccination with alternative vaccine products with and without delay. Figure showing modelled deaths (A), hospital admissions (B) and ICU admissions (C) from the original model fit (in blue) and from counterfactual scenarios (in red) with: vaccination with a Pfizer/BioNTech efficacy profile; vaccination with a Pfizer/BioNTech efficacy profile and a delay in vaccination of 2 months; vaccination with an Oxford/AstraZeneca efficacy profile; and vaccination with an Oxford/AstraZeneca efficacy profile with a delay of 2 months. Lines show the median value from 500 simulations. To facilitate comparison between scenarios, modelled deaths do not include uncertainty generated through the observation process and are therefore higher than those shown in the model fit in Figure 4.

### Trade-off between vaccination and population mobility

Finally, we investigated the trade-off between levels of vaccination coverage and population mobility on hospitalisations, ICU admissions and deaths. Here, we explore different counterfactual combinations of vaccination coverage and population mobility change to understand how much relaxation of social distancing measures vaccination could ‘buy’ in the later stages of a pandemic.

We found that, overall, changes in population mobility resulted in greater variation in disease burden than vaccination, where an increase in population mobility resulted in more additional deaths, hospitalisation and ICU admissions than a corresponding decrease in vaccination (Figure 8). For instance, our analysis suggests an increase of 20% in population mobility would result in around 10,800 extra hospital admissions, 2,400 extra ICU admissions and 800 extra deaths, while a reduction of 20% in vaccination coverage would result in around 2,000 extra hospital admissions, 600 extra ICU admissions and 200 extra deaths (Figure 8).

**Figure 8:**
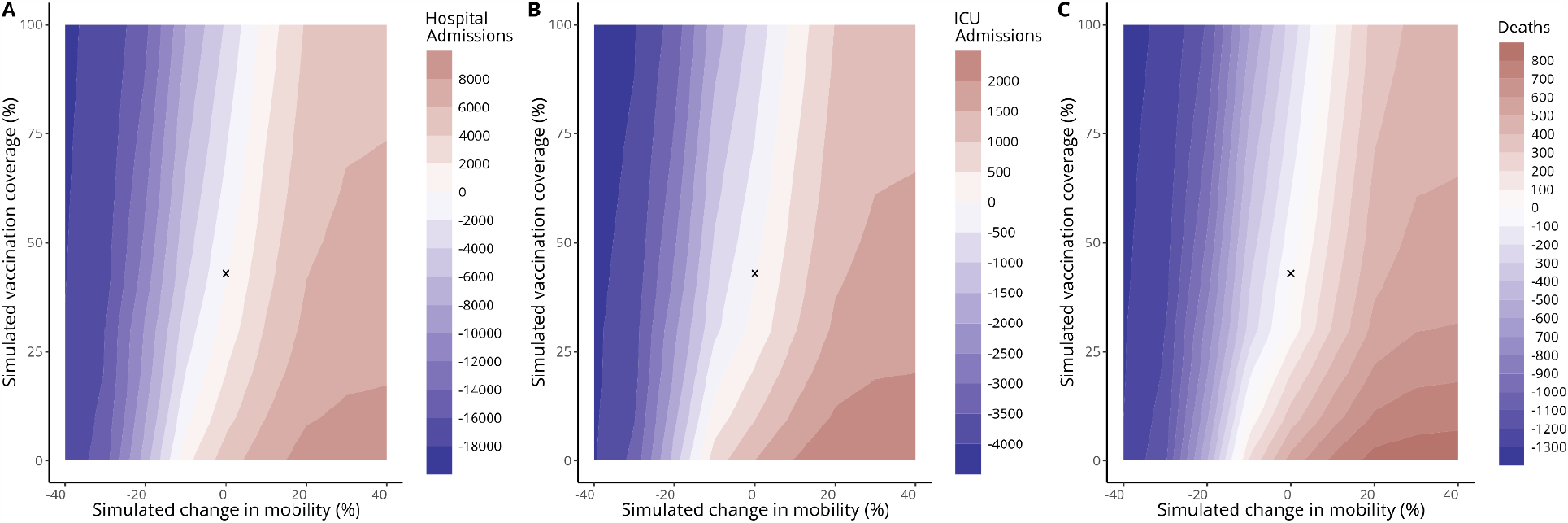
Modelled total additional hospital admissions, ICU admissions and deaths in the 6 months following the vaccination campaign launch (16th February 2021 - 16th August 2021). These contour plots show additional deaths (panel A), hospital admissions (panel B) and ICU admissions (panel C) compared to the original model fit under different levels of simulated vaccination coverage reached by 16th August 2021 (y-axis) and changes in population mobility during the 6 month period (x-axis). The actual vaccination coverage observed on 16th August 2021 is shown by a cross (vaccination coverage = 43% and mobility change = 0).

We also compared combinations of population mobility reduction and vaccination coverage that resulted in the same outcomes (looking at lines of equivalence in Figure 8). We found that in the absence of vaccination, an additional 10-20% reduction in population mobility would have been required to obtain the same hospitalisation and death outcomes seen in this period, quantifying the ‘return-to-normality’ associated with the first 6 months of vaccination in this setting.

Additionally, if population mobility had remained as measured in this period but perfect vaccination coverage had been achieved, an estimated an additional 3,740 hospital admissions (95% CrI: 3,450 – 4,030), 829 ICU admissions (95% CrI: 767 – 895) and 258 deaths (95% CrI: 224 – 297) would have been averted. This additional burden averted (going from the observed coverage of 43% to 100%) is lower than the burden averted that we estimated in the earlier ‘no vaccination’ scenario (going from 0% coverage to the observed 43% coverage, Table 2). This illustrates the importance of an age-targeted approach in reducing morbidity and mortality. Finally, a simulated change in mobility of +30% would have returned contacts in February 2021 back to (or slightly above) pre-pandemic baseline levels. This suggests that a return to baseline mobility at the beginning of the vaccination campaign would have resulted in an extra 5,130 hospital admissions (95% CrI: 6,080 – 8,480), 1,670 ICU admissions (95% CrI: 1,320 – 2,030) and 555 deaths (95% CrI: 283 – 842).

## Discussion

We used an age-structured transmission dynamic model to quantify the drivers of SARS-CoV-2 transmission in the Dominican Republic and investigated the impact of the vaccination campaign and other counterfactual vaccination scenarios. We found that despite substantial prior accumulation of post-infection immunity, the vaccination campaign had an important impact on disease burden in 2021 and was essential in enabling a return to pre-pandemic mobility levels without incurring substantial additional burden. We estimate the campaign averted 5040 hospital admissions (95% CrI: 4750 -5350), 1500 ICU admissions (95% CrI: 1420 - 1590) and 544 deaths (95% CrI: 488 - 606) in the first 6 months of the campaign.

From 2020-2021 the Dominican Republic experienced four distinct waves of SARS-CoV-2 transmission. We found that, after the initial emergence of SARS-CoV-2 in March 2020, the first wave was largely controlled by the imposition of NPIs and the associated sharp drop in contact rates. The subsequent build up of immunity in the following months maintained an estimated R_t_ of around 1 until the end of the 2020, despite the gradual increase of social contact rates from their trough. This contrasts with other settings with well-characterised transmission dynamics, such as the United Kingdom, where SARS-CoV-2 dynamics were largely driven by the imposition of NPIs before the widespread rollout of vaccination (19). The second wave from November - February 2021 was driven by a spike in contacts in December 2021, while the third wave during the summer of 2021 is best explained by gradually increasing contact rates back to pre-pandemic levels alongside the emergence of more transmissible variants, including the Mu variant. Finally, the fourth wave between September and December 2021 was driven by the Delta variant, alongside the reopening of schools and the relaxation of NPIs including curfews.

We estimated that the 43% two-dose coverage achieved by the vaccination campaign by mid-August 2021 would have offset a 10-20% increase in mobility – a proxy for social interactions – in this period, quantifying the ‘return-to-normality’ enabled by the vaccination campaign. Indeed, from July 2021 the Dominican Republic began to reopen the economy, culminating in the removal of curfew measures in October 2021 with population mobility almost returning to pre-pandemic baseline levels. This contrasts with other settings where higher levels of vaccination coverage were required to lift measures, such as the United Kingdom, which relaxed many measures in the summer of 2021 with a two-dose vaccine coverage of around 60% and population mobility still well below baseline (18,20). Many other countries were unable to lift measures before intense Omicron transmission generated substantial population immunity or until very high vaccination coverage was achieved (21). The trade-off between vaccination and population mobility on disease burden is likely to differ depending on the setting and epidemiological context. For instance, countries with lower levels of post-infection immunity would likely see greater changes in burden associated with changes in vaccination coverage. This balance will also be affected by the emergence of new variants which may be more transmissible or exhibit immune evasive properties.

There are several limitations to this analysis. During the first wave (March - September 2020), the model struggled to reproduce the observed pattern in reported deaths. This may reflect limits in testing infrastructure and COVID-19 death reporting during this period, as observed in many countries globally (22). Hospitalisation and ICU data were only publicly reported from September 2020, and deaths were probably under-reported early in the pandemic. While we partially accounted for changes in COVID-19 death reporting by allowing the infection-fatality ratio to vary over time, there remains substantial uncertainty in the modelled size and timing of the first wave.

Our modelling framework only incorporates protection from full primary vaccination with two doses and does not incorporate protection from a single dose. This would result in an underestimation of the impact of vaccination. However, as estimated protection of a single dose of Coronavac is low (particularly against the Delta strain), we do not expect this to have an important impact on our results (23).

Additionally, we assume fully vaccinated individuals are immediately afforded protection according to vaccine efficacy estimates used. We do not consider the impact of booster vaccination, which had begun in the Dominican Republic by late 2021 and we do not consider any additional benefit afforded by vaccination for individuals with post-infection immunity. Due to limited information on the introduction and epidemiological characteristics of variants introduced in early 2021, we parameterised the model for wild-type and Delta variants and modelled the effect of Mu and other VOC/VOIs in mid 2021 through a fitted sinusoidal increase in transmissibility over this period. We therefore do not capture the effect of immune evasion of Mu (or other variants such as Gamma) on the epidemic dynamics. Assuming increased transmissibility of Mu or other variants, rather than immune evasion, would result in an underestimation of the impact of the vaccination campaign, as fewer individuals would remain susceptible during vaccine roll-out and therefore able to benefit from post-vaccination rather than post-infection protection.

Our analysis untangles the complex interactions between population behaviour, the introduction of variants and changes in population immunity in the Dominican Republic, enabling us to estimate the impact of vaccination and consider other counterfactual scenarios. We quantify the impact of these epidemic drivers in a setting with high seroprevalence during vaccination rollout, providing alternative insights to much comparable modelling in high-income countries. Our conclusions are therefore likely to be relevant to many other countries that were unable to suppress transmission through NPIs prior to vaccination roll-out. We also highlight the importance of having multiple data streams available to accurately characterise transmission dynamics during an epidemic and particularly the utility of serological data in estimating population infection history. Similarly, the availability of hospital and ICU occupancy data is crucial for understanding how the relationship between infection, severe outcomes and death modulates during the epidemic due to improved treatment, the introduction of VOC/VOIs, and vaccination. Understanding these dynamics in real-time is essential to avoid potential problems such as reopening the economy too late when the population has high levels of immunity or delaying the re-imposition of NPIs when new variants emerge or contact rates increase unexpectedly. Ensuring that reliable data streams can be set-up quickly across both high income and low- and middle income countries should be a priority for future pandemic planning and preparedness.

## Data sharing

All code and data used for this analysis are available at: https://github.com/EmilieFinch/DR-covid19

## Acknowledgments

EF was supported by the Medical Research Council (MR/N013638/1); AJK was supported by Wellcome Trust (206250/Z/17/Z); RL was supported by a Royal Society Dorothy Hodgin Fellowship; EJK was supported by the US CDC (U01GH002238), CLL was supported by an Australian National Health and Medical Research Council Investigator Grant (APP1158469). CTP and RS-R are employees of the Ministry of Ministry of Health and Social Assistance, Dominican Republic.

We would like to thank the participants of the serosurvey in the Dominican Republic and the field teams that collected the data as well as the Dominican Republic Ministry of Health and Social Assistance for their support for this work. We would also like to thank Nicholas G. Davies, Rosana C. Barnard and Lloyd A.C. Chapman for helpful discussions in the completion of this work.

