## Supplementary Material for "Modelling the impact of population mobility, post-infection immunity and vaccination on SARS-CoV-2 transmission in the Dominican Republic"

### Supplementary Materials

#### Model equations

#### Table S.1: Description of model compartments

| Compartment | Description |
|---|---|
| $S_i$ | Number of susceptible individuals in age group *i*. |
| $V_i$ | Number of individuals in age group *i* protected from infection by full vaccination. Note that only susceptible individuals can enter the vaccinated compartment, and so this does not represent individuals who have both post-infection and post-vaccination immunity |
| $E_{ik}$ | Number of exposed individuals (with a latent infection) with variant *k* in age group *i* |
| $I_{sik}$ | Number of individuals with a sub-clinical (asymptomatic) infection with variant *k* in age group *i* |
| $I_{pik}$ | Number of individuals with a pre-clinical infection with variant *k* in age group *i* |
| $I_{cik}$ | Number of individuals with a clinical (symptomatic) infection with variant *k* in age group *i* |
| $R_{ik}$ | Number of individuals recovered from variant *k* in age group *i* |

$$S_i(t+1) = S_i(t) \cdot (1-\lambda_{ik}(t)) - v_i(t+1) \cdot \frac{S_i(t)}{N_i(t)} + R_{ik}(t) \cdot wn + V_i(t) \cdot wv$$

$$V_i(t+1) = v_i(t+1) \cdot \frac{S_i(t)}{N_i(t)} - V_i(t) \cdot \lambda_{ik}(t) \cdot (1-vei_k) - V_i(t) \cdot wv$$

$$E_{ik}(t+1) = E_{ik}(t) \cdot (1-\sigma) + V_i(t) \cdot \lambda_{ik}(t) \cdot (1-vei_k) \cdot (1-ved|i_k)$$

$$I_{pik}(t+1) = I_{pik}(t) \cdot \left(1-\gamma_p\right) + E_{ik}(t) \cdot \sigma \cdot y_i$$

$$I_{cik}(t+1) = I_{cik}(t) \cdot (1-\gamma_c) + I_{pik}(t) \cdot \gamma_p$$

$$I_{sik}(t+1) = I_{sik}(t) \cdot (1-\gamma_s) + E_{ik}(t) \cdot \sigma \cdot (1-y_i) + V_i(t) \cdot \lambda_{ik}(t) \cdot (1-vei_k) \cdot (ved|i_k)$$

$$R_{ik}(t+1) = R_{ik}(t) \cdot (1-wn) + I_{sik}(t) \cdot \gamma_s + I_{cik}(t) \cdot \gamma_c$$

The force of infection is given by:

$$\lambda_{ik}(t) = u_{ik} \cdot \sum_{j=1}^{J} C_{i,j,t} \cdot \frac{I_{pj}(t) + I_{cj}(t) + f \cdot I_{sj}(t)}{N_j}$$

Where $u_{ik}$ is susceptibility for age group *i* and variant *k*. *j* also depicts age group with J of 16 and *f* represents the relative infectiousness of sub-clinical (or asymptomatic) infections, which is 50%.

Hospitalisation, ICU admission and death are modelled as observation processes. Individuals enter observation processes when they mature from the $E_{ik}$ compartment based on associated delays and age-specific probabilities. Age-specific probabilities of hospitalisation, ICU admission and death are based on estimates in the literature and adjusted on the log odds scale by several fitted parameters as described in Supplementary Table S.2. The delay between infection to death is assumed to follow a gamma distribution where the mean is estimated during model fitting (and bounded between 5 and 30 days) and the shape parameter is 2.2 such that:

$$P_{death} = Gamma(mean = death\_mean, shape = 2.2)$$

$$D(t) = \sum_{i=I}^{i=I} \sum_{d=1}^{d=D} E(t-d)_i \cdot P_{death} \cdot IFR_{ik}$$

Hospitalisation and ICU admissions are estimated using the same approach with shape parameters 0.71 and 1.91 respectively.

$$P_{hosp} = Gamma(mean = hosp\_mean, shape = 0.71)$$

$$Hosp(t) = \sum_{i=I}^{i=I} \sum_{d=1}^{d=D} E(t-d)_i \cdot P_{hosp} \cdot ISR_{ik}$$

$$P_{ICU} = Gamma(mean = icu\_mean, shape = 1.91)$$

$$ICU(t) = \sum_{i=I}^{i=I} \sum_{d=1}^{d=D} E(t-d)_i \cdot P_{ICU} \cdot ICR_{ik}$$

The full likelihood is the sum of the likelihoods listed below.

$$Y_{Hosp}(t) \sim NegBin(X_{Hosp}(t), \kappa_{Hosp})$$

$$L_{Hosp} = \sum P_{NegBin}(Y_{Hosp}(t) | X_{Hosp}(t), \kappa_{Hosp})$$

$$Y_{ICU}(t) \sim NegBin(X_{ICU}(t), \kappa_{ICU})$$

$$L_{ICU} = \sum P_{NegBin}(Y_{ICU}(t) | X_{ICU}(t), \kappa_{ICU})$$

$$Y_{Deaths}(t) \sim NegBin(X_{Deaths}(t), \kappa_{Deaths})$$

$$L_{Deaths} = \sum P_{NegBin}(Y_{Deaths}(t) | X_{Deaths}(t), \kappa_{Deaths})$$

$$Y_{Sero}(t) \sim SkewNorm(X_{Sero}(t), \xi_{Sero}, \omega_{Sero}, \alpha_{Sero})$$

$$L_{Sero} = \sum_{s=1}^{S} P_{SkewNorm}(Y_{Sero}(t) | X_{Sero}(t), \xi_{Sero}, \omega_{Sero}, \alpha_{Sero})$$

### Table S.2: Fixed model parameters

| Parameter | Description | Value | Reference |
|---|---|---|---|
| tS | Start date of wild-type SARS-CoV-2 epidemic in days after 1 Jan 2020 | 20 | Determines the date which seeding begins (28) |
| dE | Latent period (E to Ip and E to Is in days) | ~gamma($\mu$ = 2. 5, $k$ = 2. 5) | 2.5 so that the incubation period (latent period plus period of preclinical infectiousness is 5 days). Lauer et al, 2020 (29) |
| dP | Duration of pre-clinical infectiousness | ~gamma($\mu$ = 2. 5, $k$ = 4) | Assumed to be half the duration of total infectiousness in clinically-infected individuals (30) |
| dC | Duration of clinical infectiousness | ~gamma($\mu$ = 2. 5, $k$ = 4) | Infectious period set to 5 days to result in a serial interval of approximately 6 days (31-33) |
| dS | Duration of subclinical infectiousness | ~gamma($\mu$ = 5. 0, $k$ = 4) | Assumed to be the same duration as total infectious period for clinical cases, including preclinical transmission |
| yi | Probability of clinical symptoms given infection for age group *i* | Estimated from case distribution across 6 countries | (34) |
| f | Relative infectiousness of subclinical cases | 50% | Assumed (24, 34) |
| Cij | Number of age-j individuals contacted by an age-i-individual per day, prior to changes in mobility | Dominican Republic specific contact matrix | (35) |
| Ni | Number of age-i individuals | From demographic data | |
| Δt | Time step for discrete-time simulation | 0.25 days | |
| P.death | Infection-fatality ratio by age | | Using estimates based on age-specific death data from 45 countries and 22 seroprevalence surveys from O'Driscoll et al (36) |
| P.hosp | Infection-severe ratio by age (infections resulting in hospitalisation) | | Using estimates based on on multi-country serology studies in Herrera-Esposito et al (37) |
| P.critical | Infection-critical ratio (infections resulting in admission to ICU) | | Using estimates based on multi-country serology studies in Herrera-Esposito et al (37) |
| extra_voc_takeoff | Dates during which transmissibility increases according to a logistic growth function | 8th February - 5th April | Informed by GISAID sequence data (16) |

### Table S.3: Fitted parameters and prior distributions

| Parameter | Description | Prior | Note |
|---|---|---|---|
| u | Basic susceptibility to infection | ~normal(0.09, 0.02) ≥ 0.05 and ≤ 0.2 | Determines the basic reproduction number, $R_0$ |
| death_mean | Mean delay in days from start of infectious period to death | ~normal(15, 2) ≥5 and ≤30 | Delay is assumed to follow a gamma distribution with shape parameter 2.2. Prior and shape of distribution informed by analysis of CO-CIN data (38) |
| hosp_admission | Mean delay in days from start of infectious period to hospitalisation | ~normal(8, 1) ≥4 and ≤20 | Delay is assumed to follow a gamma distribution with shape parameter 0.71. Prior and shape of distribution informed by analysis of CO-CIN data (38) |
| icu_admission | Mean delay in days from start of infectious period to ICU admission | ~normal(12.5, 1) ≥8 and ≤14 | Delay is assumed to follow a gamma distribution with shape parameter 1.91. Prior and shape of distribution informed by analysis of CO-CIN data (38) |
| cfr_rlo<br>cfr_rlo2<br>cfr_rlo3 | Relative log-odds of death from COVID-19 for different time periods | ~normal(0, 0.1) ≥-2 and ≤2 | Age-specific case fatality rates based on O'Drisoll et al (36). This was adjusted by cfr_flo, cfr_rlo2 and cfr_rlo3 over time |
| hosp_rlo<br>hosp_rlo2 | Log-odds of hospital admission given infection derived from Herrera-Eposito et al. relative to age-specific probabilities of hospital admission in first half of 2020 | ~normal(0, 0.1) ≥-2 and ≤2 | Age-specific probabilities of hospitalisation are based on Herrera-Esposito et al (37) then adjusted based on the icu_rlo and icu_rlo2 parameters. icu_rlo applies for the first half of 2020 while icu_rlo2 applies for the second half of 2020 into 2021. |
| icu_rlo<br>icu_rlo2 | Log-odds of ICU admission relative to age-specific probabilities of ICU admission in first half of 2020 | ~normal(0, 0.1) ≥-2 and ≤2 | Age-specific probabilities of ICU admission are based on Herrera-Esposito et al (37). These are then adjusted based on the icu_rlo and icu_rlo2 parameters. icu_rlo applies for the first half of 2020 while icu_rlo2 applies for the second half of 2020 into 2021. |
| disp_deaths<br>disp_hosp_inc<br>disp_hosp_prev<br>disp_icu_prev | Negative binomial dispersion for deaths, hospital incidence (admissions), hospital prevalence (beds occupied), and ICU prevalence | ~exponential(10) | We estimate the size parameter for negative binomial likelihood functions of deaths, hospital incidence, hospital prevalence and ICU prevalence, where size = $1/(\text{disp}^2)$ |
| contact_adj_a<br>contact_adj_b | Parameter determining the weighting given to comix-adjusted contacts vs baseline contacts from Google mobility data. contact_adj_a applies in 2020 and contact_adj_b applies in 2021 | ~beta(15, 1) | To account for differences in the relationship between Google mobility data and contact patterns between the UK and the Dominican Republic, we allow modelled contacts to be scaled towards pre-pandemic baseline values through fitted parameters contact_adj_a and contact_adj_b. |
| extra_voc_relu | Relative transmissibility of Mu (and other VOI in mid 2021) | ~lognormal(0.4, 0.1) | Prior centred around estimated transmission advantage of VOIs over WT of 1.5 |
| v2_relu | Relative transmissibility of Delta variant | ~lognormal(0.92, 0.1) | Prior centred around estimated transmission advantage of Delta over WT of 2.5. (25) |
| v2_when | Date of introduction of Delta variant in days after 1st Jan 2020 | ~uniform(486, 517) | On this date, ten random individuals contract B.1.617.2 (Delta). We use a uniform prior between 1st May 2021 - 1st June 2021 based on sequence data available for the Dominican Republic (see Figure 2) |
| v2_hosp_rlo<br>v2_icu_rlo<br>v2_cfr_rlo | Relative log-odds of hospitalisation, ICU admission and death for Delta compared to pre-existing variants | ~normal(0, 0.1) ≥-4 and ≤4 | Vague priors |

### Table S.4: Vaccine efficacy parameters

| Description | Value | Reference |
|---|---|---|
| Overall efficacy against infection with ancestral variant | 0.67 | Assumed to be the same as efficacy against disease. |
| Overall efficacy against disease with ancestral variant | 0.67 | Imai et al (Table 2) (23) |
| Overall efficacy against infection with Delta variant | 0.39 | Assumed to be the same as efficacy against disease. |
| Overall efficacy against disease with Delta variant | 0.39 | Wu et al 2022 (Table 3) (39) |

### Table S.5: Waning parameters

| Description | Default value (central waning) |
|---|---|
| Rate of waning from the recovered compartment to the susceptible compartment for all strains | log(0.85)/-365,corresponding to exponential waning with a 15% loss of protection after 1 year |
| Rate of waning from the vaccinated compartment to the susceptible compartment | log(0.6)/-182.5, corresponding to exponential waning with a 40% loss of protection after 6 months. Based on Cerqueira-Silva et al (40) |

**Figure S.1: Posterior estimates of fitted parameter**

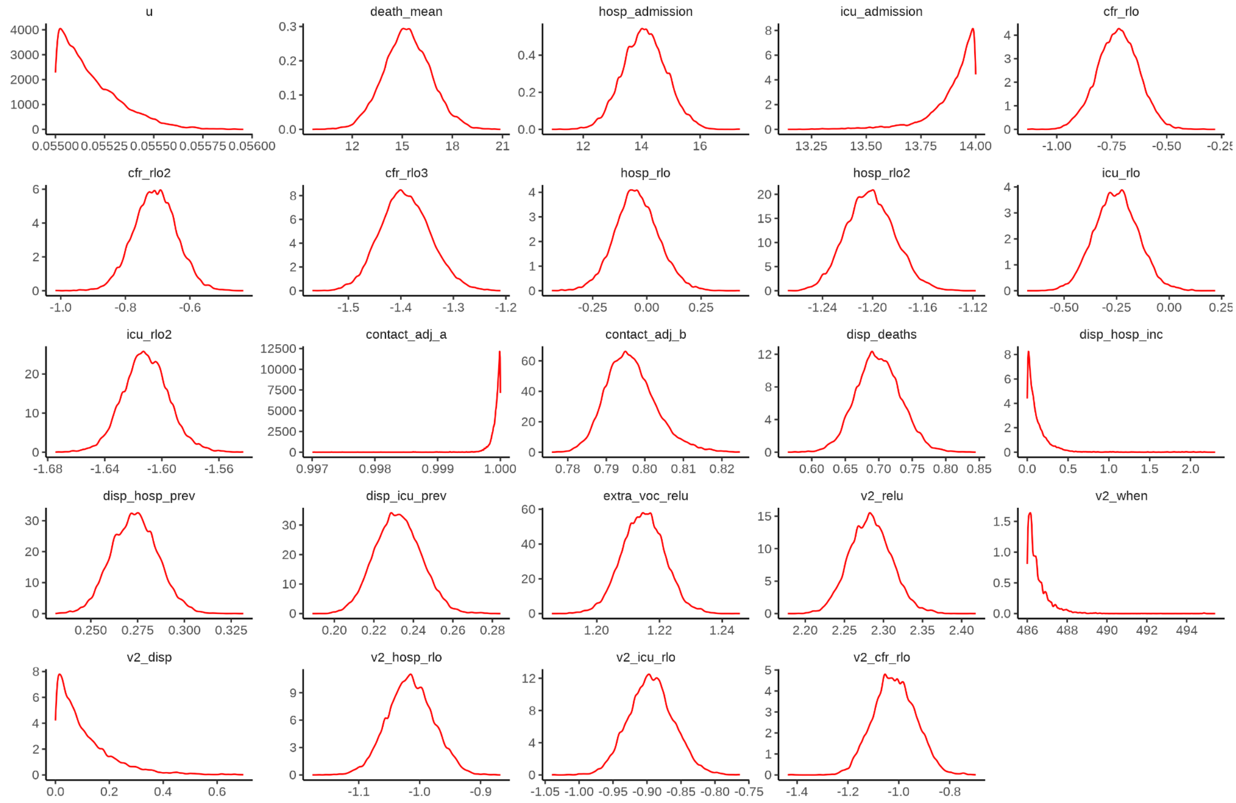


**Table S.6: Vaccine efficacy parameters for two doses of Pfizer and AstraZeneca vaccines against WT and Delta variants.**

| Description | Value |
|---|---|
| Pfizer overall efficacy against infection with WT | 0.85 |
| Pfizer overall efficacy against disease with WT | 0.9 |
| Pfizer overall efficacy against infection with Delta variant | 0.8 |
| Pfizer overall efficacy against disease with Delta variant | 0.81 |
| AZ overall efficacy against infection with WT | 0.75 |
| AZ overall efficacy against disease with WT | 0.8 |
| AZ overall efficacy against infection with Delta variant | 0.63 |
| AZ overall efficacy against disease with Delta variant | 0.65 |

Table adapted from Supplementary Table 6 and 7 in Barnard et al 2022 (19)

**Table S.7: Sensitivity analysis: estimated deaths, hospital admissions and ICU admissions under different waning assumptions**

| Scenario | Additional hospital admissions in next 6 months | Additional hospital admissions in next 10 months | Additional ICU admissions in next 6 months | Additional ICU admissions in next 10 months | Additional deaths in next 6 months | Additional deaths in next 10 months |
|---|---|---|---|---|---|---|
| No vaccination (central waning) | 5040 (4750 – 5350) | 5150 (4770 – 5540) | 1500 (1420 – 1590) | 1530 (1420 – 1640) | 544 (488 – 606) | 554 (487 – 627) |
| No vaccination (high waning) | 4490 (4250 – 4780) | 4350 (4030 – 4720) | 1330 (1250 – 1400) | 1260 (1170 – 1360) | 472 (421 – 536) | 450 (382 – 528) |
| No vaccination (low waning) | 5450 (5150 – 5820) | 5790 (5390 – 6250) | 1630 (1540 – 1730) | 1730 (1610 – 1860) | 577 (514 – 653) | 611 (536 – 701) |
| Pfizer instead (central waning) | -1830 (-1940 – -1730) | -3410 (-3790 – -3070) | -542 (-573 – -512) | -1020 (-1120 – -921) | -193 (-215 – -174) | -340 (-414 – -273) |
| Pfizer instead (high waning) | -1650 (-1750 – -1560) | -2480 (-2820 – -2160) | -483 (-509 – -457) | -725 (-820 – -633) | -171 (-194 – -152) | -255 (-346 – -171) |
| Pfizer instead (low waning) | -1970 (-2100 – -1870) | -4120 (-4530 – -3770) | -581 (-618 – -553) | -1230 (-1340 – -1140) | -203 (-230 – -181) | -395 (-484 – -322) |
| Pfizer delayed (central waning) | 2980 (2760 – 3210) | 124 (-364 – 596) | 811 (753 – 870) | -65 (-188 – 60) | 283 (236 – 332) | 17 (-89 – 114) |
| Pfizer delayed (high waning) | 2540 (2350 – 2760) | 279 (-179 – 743) | 672 (619 – 724) | -14 (-138 – 105) | 227 (180 – 278) | -7 (-140 – 107) |
| Pfizer delayed (low waning) | 3270 (3030 – 3550) | 86 (-437 – 610) | 894 (837 – 967) | -77 (-204 – 60) | 306 (255 – 368) | 22 (-99 – 135) |
| AZ instead (central waning) | -1040 (-1100 – -981) | -1920 (-2150 – -1700) | -307 (-325 – -290) | -577 (-642 – -515) | -109 (-122 – -98) | -193 (-240 – -150) |
| AZ instead (high waning) | -923 (-979 – -874) | -1370 (-1580 – -1170) | -271 (-286 – -257) | -404 (-463 – -346) | -96 (-109 – -86) | -142 (-199 – -89) |
| AZ instead (low waning) | 1120 (-1200 – -1060) | -2350 (-2610 – -2120) | -331 (-352 – -315) | -708 (-778 – -647) | -115 (-131 – -103) | -227 (-284 – -179) |
| AZ delayed (central waning) | 3220 (3000 – 3450) | 1090 (654 – 1520) | 892 (834 – 952) | 234 (124 – 348) | 313 (267 – 361) | 113 (20 – 200) |
| AZ delayed (high waning) | 2770 (2580 – 2990) | 1070 (661 – 1490) | 748 (696 – 800) | 228 (117 – 336) | 255 (211 – 305) | 77 (-37 – 179) |
| AZ delayed (low waning) | 3520 (3290 – 3810) | 1160 (695 – 1640) | 980 (921 – 1050) | 254 (141 – 381) | 338 (287 – 399) | 126 (21 – 228) |

### Table S.8: Sensitivity analysis: description of waning assumptions

| Waning scenario | Description | Default value (central waning) |
|---|---|---|
| Central | Rate of waning from the recovered compartment to the susceptible compartment for all strains | log(0.85)/-365,corresponding to exponential waning with a 15% loss of protection after 1 year |
| | Rate of waning from the vaccinated compartment to the susceptible compartment | log(0.6)/-182.5, corresponding to exponential waning with a 40% loss of protection after 6 months. Based on Cerqueira-Silva et al (33) |
| High | Rate of waning from the recovered compartment to the susceptible compartment for all strains | log(0.85)/-182.5,corresponding to exponential waning with a 15% loss of protection after 6 months |
| | Rate of waning from the vaccinated compartment to the susceptible compartment | log(0.6)/-91.25 ,corresponding to exponential waning with a 40% loss of protection after 3 months |
| Low | Rate of waning from the recovered compartment to the susceptible compartment for all strains | log(0.85)/-730,corresponding to exponential waning with a 15% loss of protection after 2 years |
| | Rate of waning from the vaccinated compartment to the susceptible compartment | log(0.84)/-120 ,corresponding to exponential waning with a 16% loss of protection after 4 months. |

**Figure S.2: Modelled hospitalisations, deaths, and proportion previously infected by age group**

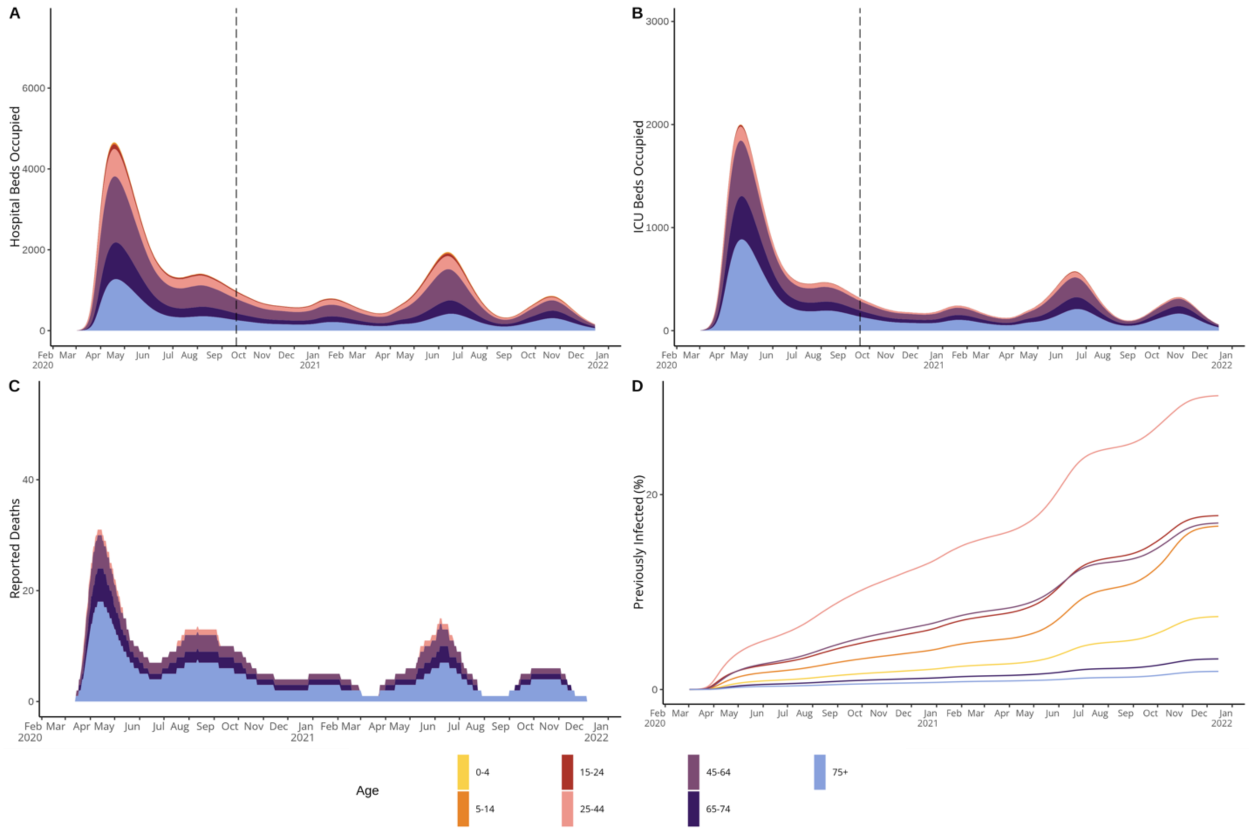

### Figure S.3: Modelled immune status of the population by age group

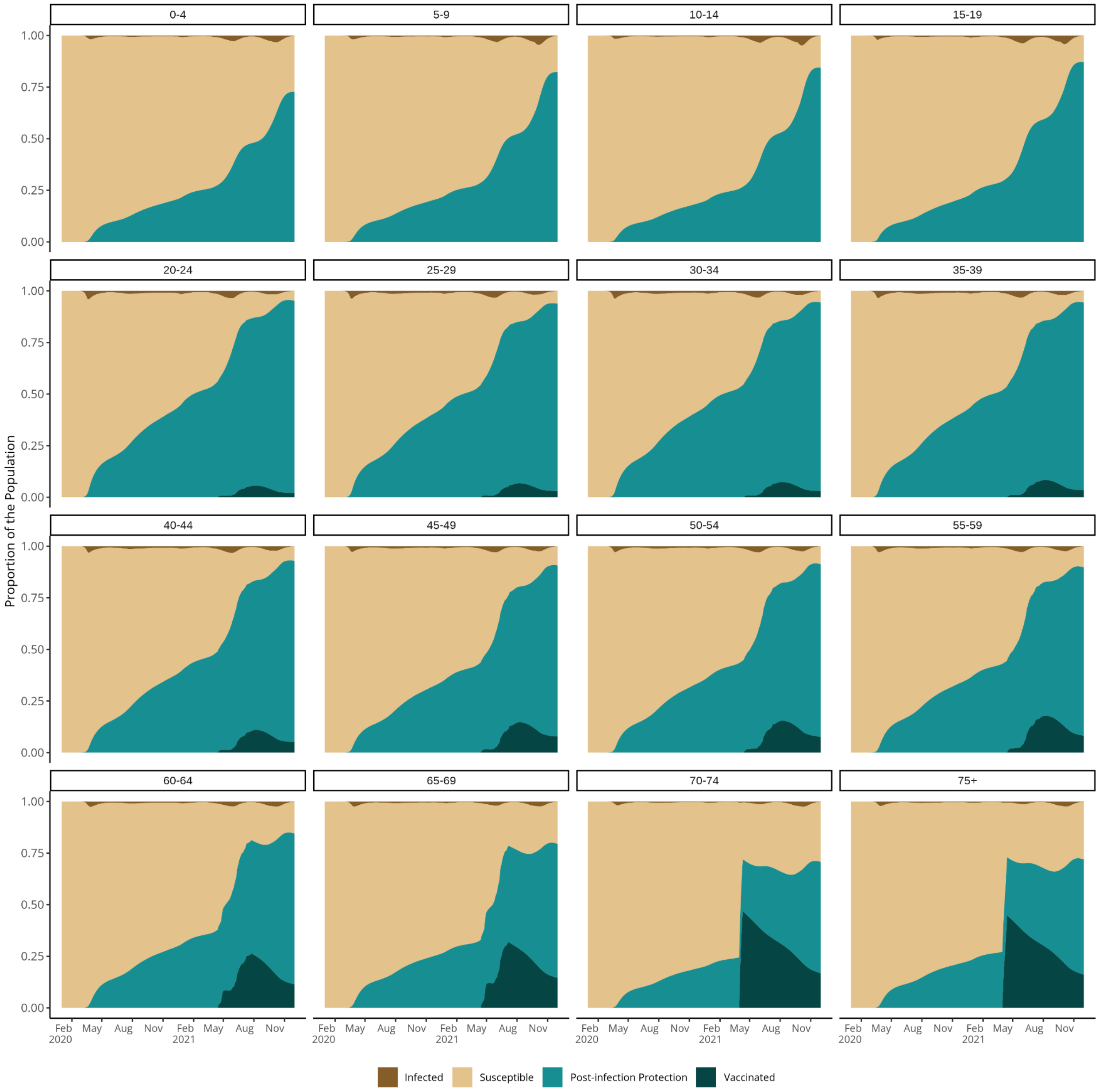

**Table S.9 Eligibility of age groups for vaccination in the Dominican Republic**

| Phase | Date | Age group |
| --- | --- | --- |
| Fase IC | 25/02/2021 | > 70 |
| Fase ID | 24/04/2021 | > 60 |
| Fase II | 03/05/2021 | > 50 |
| Fase III | 10/05/2021 | > 18 |
